# Future Pandemics: AI-Designed Diagnostic Assays for Detection of Andes Orthohantavirus (ANDV) Associated with the 2026 MV Hondius Outbreak

**DOI:** 10.64898/2026.05.26.26354101

**Authors:** John MacSharry, Alberto Tonda, Alejandro Lopez-Rincon

## Abstract

Andes orthohantavirus (ANDV), the primary etiological agent of hantavirus pulmonary syndrome (HPS) in South America, is uniquely capable of limited human-to-human transmission, posing a significant challenge for outbreak control. Recent events, including the 2018–2019 Epuyén outbreak and the 2026 MV Hondius incident, underscore the need for rapid, lineage-specific molecular diagnostics. In this study, we present an artificial intelligence (AI)-driven framework for the design of diagnostic primers targeting the S genomic segment of the Epuyén lineage. Using an evolutionary algorithm integrated with thermodynamic evaluation via Primer3Plus, candidate primers were optimized to maximize classification accuracy while satisfying stringent biochemical constraints. The resulting primer set enables amplification of lineage-specific regions suitable for molecular characterization and surveillance. In silico validation demonstrates that the proposed primers achieve perfect discrimination between 2026 outbreak sequences and other ANDV variants. Furthermore, *in silico* comparison with standard protocol-based primers reveals substantially reduced sensitivity and specificity in the latter, highlighting the limitations of static diagnostic designs when applied to evolving viral populations. Overall, this work demonstrates that AI-assisted primer design provides a robust and adaptable strategy to improve viral detection, enhance outbreak tracking, and support timely public health interventions. Integrating computational optimization into diagnostic development is essential for strengthening preparedness against emerging zoonotic threats.

## 1 Introduction

Andes orthohantavirus (ANDV) is a zoonotic pathogen endemic to South America and is the principal causative agent of hantavirus pulmonary syndrome (HPS) in the region. Unlike most hantaviruses, which are transmitted exclusively from rodents to humans, ANDV is notable for its capacity to support limited human-to-human transmission under specific conditions, particularly among close contacts^1^. Specialized literature reports that the human-to-human spread of ANDV can occur through both the respiratory airway and saliva^2^. This distinctive feature has drawn significant attention to specific viral lineages associated with outbreaks in Argentina and Chile.

Human-derived isolates of ANDV are relatively scarce, primarily due to the difficulty of isolating the virus in cell culture following the development of neutralizing antibodies^3^. Nevertheless, several isolates obtained from human infections have contributed substantially to our understanding of viral diversity, transmission dynamics, and pathogenicity. Among these, the Epuyén lineage, associated with the 2018–2019 outbreak in Argentina, is of particular relevance due to well-documented chains of person-to-person transmission^4,5^.

The Epuyén outbreak represents one of the strongest pieces of evidence for human-to-human transmission in hantaviruses and has been associated with clustered infections exhibiting epidemiological characteristics consistent with short transmission chains. For this reason, the Epuyén lineage is considered a critical target for molecular detection and surveillance, particularly in scenarios where human-to-human transmission is suspected^5^. More recently, a multi-country outbreak of Andes orthohantavirus (ANDV) in 2026, linked to the MV *Hondius* cruise expedition, has raised additional concerns regarding transmission dynamics in confined and highly mobile populations. This event, involving confirmed cases and fatalities across international travelers, underscores the potential for rapid dissemination of ANDV beyond endemic regions and highlights the importance of developing sensitive and lineage-aware diagnostic tools for early detection and outbreak control^6^.

A representative reference sequence for this lineage is available in GenBank under accession number MN258238, corresponding to the S genomic segment of the Andes virus isolate Epuyén/18-19^7^. This segment encodes the nucleocapsid (N) protein and contains an open reading frame of approximately 1,293 nucleotides, consistent with other Andes virus S segments. Additional related sequences, such as MN258239, have been reported from the same outbreak and are frequently used for phylogenetic comparisons within this clade. Genomically, the Epuyén lineage is characterized by its relative stability and its close relationship to the Epilink/96 strain, which represents the first documented case of person-to-person hantavirus transmission in Argentina^8^.

Genomic characterization of ANDV isolates has revealed considerable diversity across lineages, with specific variants linked to distinct epidemiological and biological features^8^. Although not all human-derived samples are suitable for laboratory propagation, numerous sequences obtained directly from clinical cases have enabled detailed comparative analyses, shedding light on the molecular determinants of virulence and transmissibility^9^.

The genome of ANDV consists of three negative-sense, single-stranded RNA segments: the large (L), medium (M), and small (S) segments^10^. The L segment encodes the RNA-dependent RNA polymerase responsible for viral replication, while the M segment encodes glycoproteins involved in host cell entry. The S segment encodes the nucleocapsid protein, which plays a central role in viral assembly and viral RNA encapsidation, and in ANDV additionally encodes a non-structural protein implicated in immune evasion^11^. Due to its functional importance and relative conservation, the S segment is frequently targeted in molecular studies, including primer design for detection and characterization of specific viral lineages^12^.

In this context, the identification of lineage-specific genomic regions provides an opportunity to design targeted molecular assays capable of distinguishing between ANDV variants^8^. Such approaches are particularly relevant for tracking outbreak-associated strains, such as the Epuyén lineage, and for investigating whether ongoing outbreaks, including those reported in highly confined environments, may involve variants with enhanced human-to-human transmission potential.

## 2 Methods

### 2.1 Dataset

A total of 214 Andes virus (hantavirus) sequences were obtained from^13^, of which 210 were collected in 2024 or earlier, and 4 correspond to the 2026 outbreak. A binary classification problem was defined, where 2026 sequences were labeled as 1 and all remaining sequences as 0.

### 2.2 Primer Evaluation and Optimization

Candidate primers were defined as fixed-length subsequences of *ℓ* = 21 nucleotides extracted from the dataset. Each candidate solution is encoded as a pair *g* = (*k, p*), where *k* is the index of the sequence and *p* is the starting position (Fig. 1).

**Figure 1.**
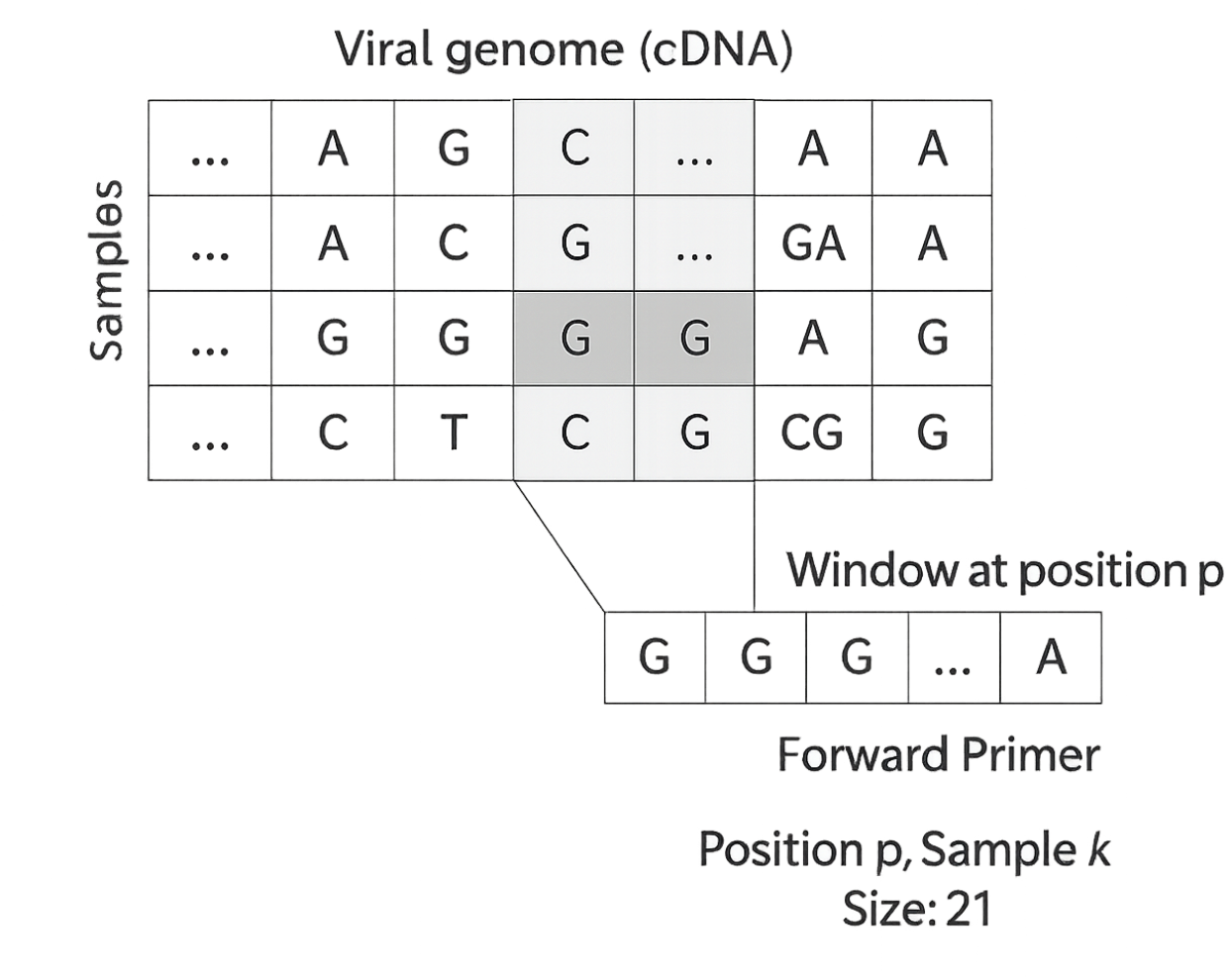
Internal encoding of a candidate primer during the EA run. A candidate sequence is identified as a target 21 bp sequence starting at position *p*, inside sample *k*, with *p* and *k* being the two integer variables optimized by the EA.

The resulting primer sequence is:

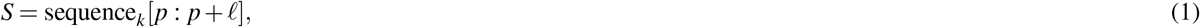

subject to the constraint:

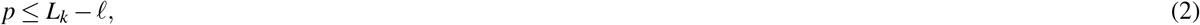

where *L*_*k*_ is the length of sequence *k*.

Primer candidates were evaluated using a cost function combining classification performance, sequence validity, and thermodynamic properties:

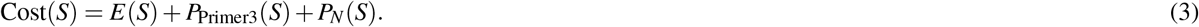

The classification error is defined as:

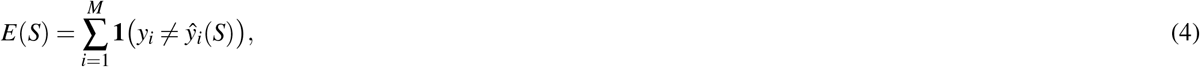

where *y*_*i*_ is the true label of sequence *i*, and the predicted label is:

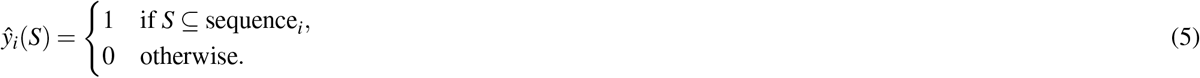

Ambiguous nucleotides were strongly penalized:

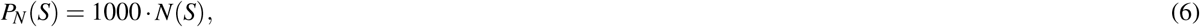

where *N*(*S*) is the number of ambiguous bases (“N”) in the primer sequence.

Thermodynamic and compositional properties were evaluated using a Primer3^14^-inspired penalty function:

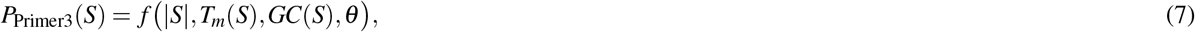

where *θ* is the set of input parameters:

- the primer length (`length`)
- melting temperature (`tm`)
- GC content (`gc`, optional)
- positional penalty (`posPenalty`)
- optimal values (`sizeOpt`, `tmOpt`, `gcOpt`),
- and weighting coefficients controlling penalties for deviations.

The melting temperature *T*_*m*_(*S*) is computed using the nearest-neighbor model:

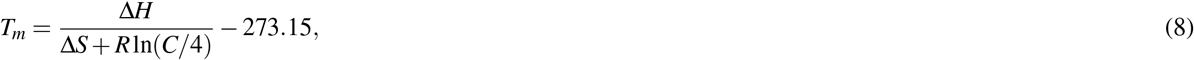

and GC content is defined as:

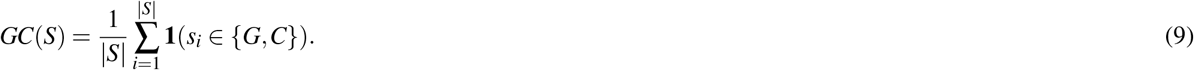

The function *f* (·) penalizes deviations from optimal ranges of primer length, melting temperature, GC content, and positional constraints. Lower cost values indicate better primer candidates.

### 2.3 Evolutionary Optimization

Primer optimization was performed using an evolutionary algorithm (EA)^15,16^. The initial population consisted of *N* = 200 individuals generated by sampling:

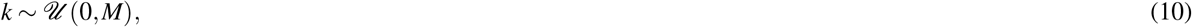

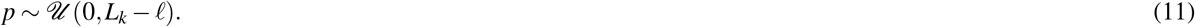

Selection was performed using tournament selection with tournament size 2. Crossover between two parents *g*^(1)^ = (*k*_1_, *p*_1_) and *g*^(2)^ = (*k*_2_, *p*_2_) produced offspring:

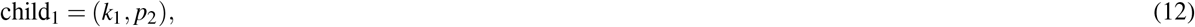

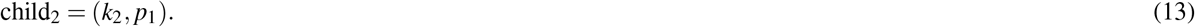

Mutation was applied independently to each component with probability *µ* = 0.15:

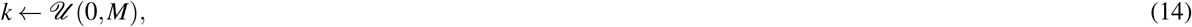

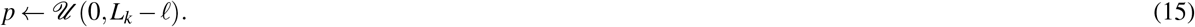

The population was updated using generational replacement, and the algorithm was run for 100 generations. Due to the stochastic nature of the EA, the optimization was repeated 20 independent times. Across runs, highly similar primer sequences were consistently identified, with the best candidate recurring in the majority of executions, indicating robustness of the method.

The optimal primer was defined as:

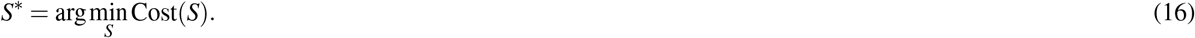

### 2.4 Reverse Sequence Primer Set Design

Following identification of the optimal forward primer, the complete primer set is designed using Primer3Plus^17^. The sequence **PP_006WBLH** (*ANDV/Switzerland/Hu-3337/2026*) was used as the reference template to design the reverse primer and internal oligonucleotide.

Standard design constraints were applied: primer length between 18-25 nucleotides, melting temperature between 58–62^◦^C, and GC content between 40–60%, consistent with Primer3 guidelines. Candidate primers were further evaluated to avoid secondary structures such as hairpins and primer dimer formation. This combined approach enabled the design of a primer set specifically targeting the Andes virus sequence within the S segment.

## 3 Results and Discussion

### 3.1 ANDV 2026 Specific Primer Design Results

We ran the EA system for 20 evaluations, the average of all the runs is shown in Fig. 2 . From the results, 4 sequences fell on the sequences of the 2026 ANDV MV Hondius Outbreak. Nevertheless, 3 subsequences presented hairpin secondary structures, which they can reduce amplification efficiency and accuracy^18^. At the end of the *in-silico* experiments, we obtained a candidate forward primer sequence “**5’–GCG TTG TAT GTT GCA GGG GTA–3’**”.

**Figure 2.**
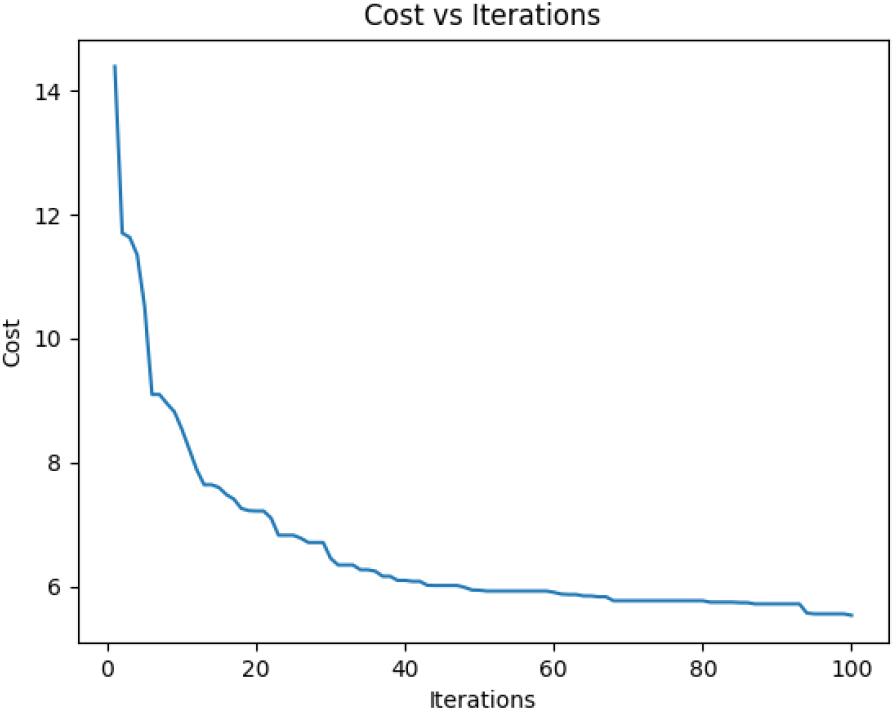
Average of the cost function for the best candidate sequence identified at each iteration of the EA optimization. The decrease in value over the course of the iteration proves that the algorithm is finding better solutions over time.

### 3.2 Primer Set Sequences

Using sequence **PP_006WBLH**, three primer sets were designed and evaluated based on their thermodynamic properties, secondary structure formation, and suitability for quantitative PCR (qPCR). A comparative summary is provided in Table 1.

**Table 1.**
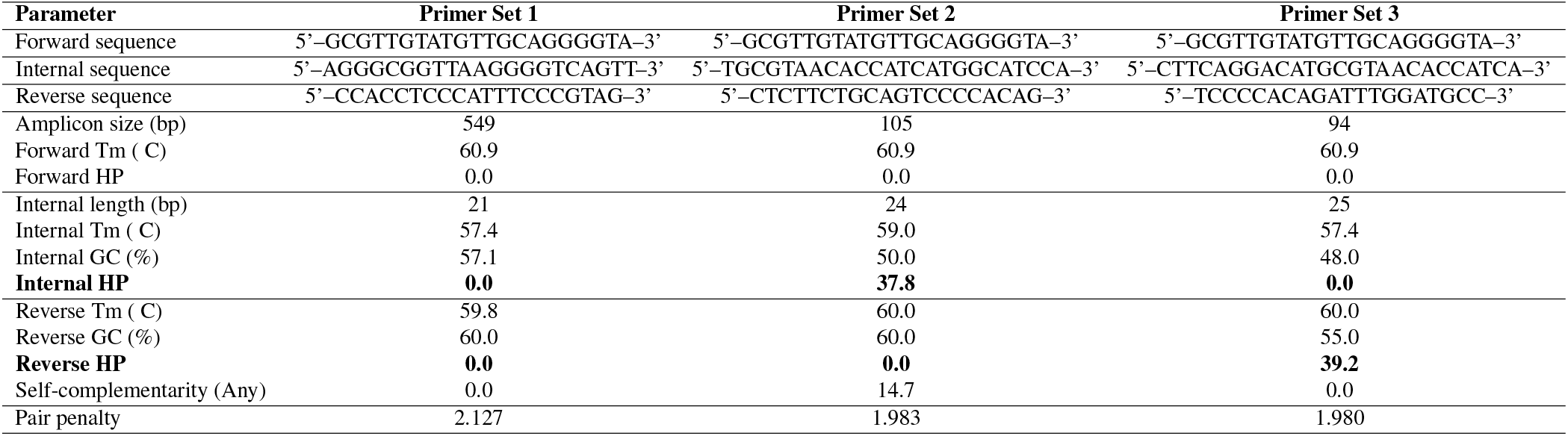
Comparison of 3 primer sets including sequences and structural properties, generated with the EA system.

Primer set 1 exhibited optimal structural characteristics, with no detectable hairpin formation or self-complementarity in any of its components. However, the resulting amplicon size (549 bp) is substantially larger than the recommended range for qPCR applications, which typically favors shorter fragments for efficient amplification. Therefore, despite its favorable structural profile, this primer set is not suitable for quantitative analysis. Nevertheless, it can be used downstream for phylogenetic tracking and molecular characterization or long-read sequencing platforms such as Nanopore^19^.

Primer set 2 produced an amplicon of 105 bp, which falls within the ideal size range for qPCR. The melting temperatures and GC content of the primers were also within acceptable limits. Nevertheless, a strong hairpin structure was identified in the internal oligo (HP = 37.8), which is likely to hinder proper hybridization and reduce assay efficiency. In addition, increased self-complementarity was observed in the reverse primer, raising the risk of non-specific interactions and reduced specificity.

Primer set 3 generated an amplicon of 94 bp, which is optimal for qPCR applications. The internal oligo in this set showed no evidence of secondary structure formation, representing a clear improvement over primer set 2. However, the reverse primer exhibited a strong hairpin structure (HP = 39.2), which may interfere with primer annealing and extension during amplification.

Overall, primer set 3 represents the most promising design due to its optimal amplicon size and improved internal oligo properties. However, further optimization of the reverse primer is required to eliminate the hairpin structure and ensure efficient and reliable qPCR performance.

### 3.3 Comparison to existing Primers Set

To evaluate the performance of two diagnostic primer sets for Hantavirus 2026 MV Hondius Outbreak detection, a confusion matrix analysis was conducted using the dataset comprising 4 2026-positive and 210 other-Hantavirus samples. In addition, we tested with 5 extra sequences of the background set from the 2026 outbreak to validate further with the same results^20^.

Figure 3 summarizes the classification outcomes. The forward sequence and primer set developed in this study achieved perfect discrimination between classes, correctly identifying all positive and negative samples, yielding both sensitivity and specificity equal to >99%. In contrast, the forward sequence and primer set recommended by the standard protocol^21^ exhibited markedly inferior performance. Although it failed to detect any of the 2026-positive samples (0/4 true positives), it also produced a substantial number of false positives among the negative samples (86/210), resulting in reduced specificity, *in silico*. The reason is that the sequence **GCAGCTGTGTCTACATTGGAGAC** has a SNP (Single Nucleotide Polymorphism) in comparison to the sequences 2026-positive **GCAGCTGTGTCTACACTGGAGAC**. This type of internal mismatch is unlikely to cause false-positive results, but it can slightly reduce amplification efficiency, which in low viral load samples could increase the risk of false-negative or weak detection signals^22^.

**Figure 3.**
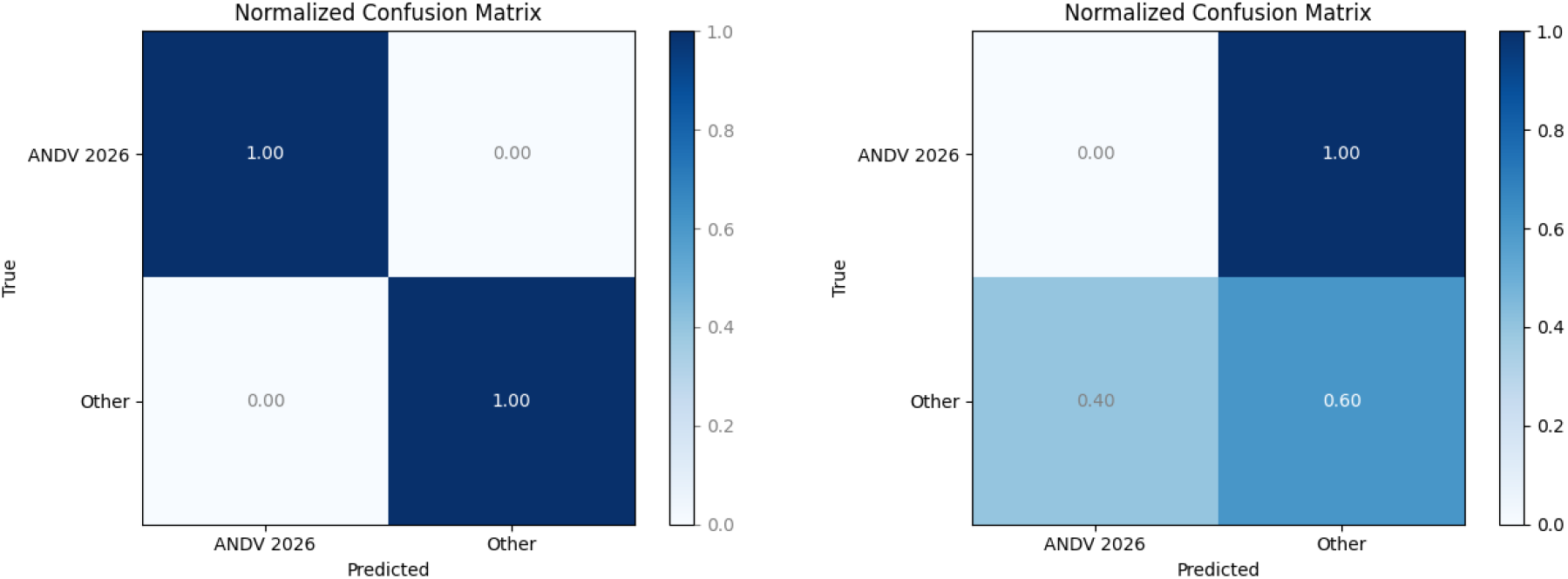
Confusion matrices for the two diagnostic primer sets evaluated for Hantavirus detection during the 2026 outbreak. The proposed primer set demonstrates perfect classification performance, correctly identifying all 2026-positive (n = 4) and other-Hantavirus (n = 210) samples without false positives or false negatives. In contrast, the protocol-recommended primer set fails to detect any positive samples (0/4 true positives) and misclassifies a substantial proportion of negative samples as positive (false positives, n = 86), resulting in both reduced sensitivity and specificity.

These results highlight a critical difference in diagnostic performance between the two approaches. While the protocol-recommended primers were not specifically optimized for the circulating Hantavirus strains associated with the 2026 outbreak, the proposed primer set demonstrates superior adaptability and robustness, effectively capturing sequence variability and improving detection accuracy. This suggests that data-driven primer design strategies can significantly enhance diagnostic reliability, particularly in the context of emerging or rapidly evolving viral outbreaks.

## Data Availability

The code for the EA system is available in
https://github.com/steppenwolf0/primersHantavirus} The used sequences are available at https://doi.org/10.62599/PP_SS_1915.1. The sequences PP_006XDJH, PP_006XDHK, PP_006XBKH} and PP_006WBLH for the focal set were used with consent of the authors.

https://github.com/steppenwolf0/primersHantavirus

## Data Availabilty

The code for the EA system is available in https://github.com/steppenwolf0/primersHantavirus. The used sequences are available at^20^. The sequences **PP_006XDJH, PP_006XDHK, PP_006XBKH** and **PP_006WBLH** for the focal set were used with consent of the authors.

